# Determinants of One-Year Mortality After Hip Fracture in U.S. Older Adults: A Socio-Ecological Systematic Review and Meta-Analysis

**DOI:** 10.64898/2026.02.10.26346053

**Authors:** Oluwaseun Adeyemi, Dowin Boatright, Joshua Chodosh

## Abstract

**Background:** Hip fracture remains a leading cause of morbidity and mortality among older adults in the United States. The aim of this systematic and meta-analytical review is to synthesize available evidence on predictors of one-year mortality following hip fracture among older adults, guided by a socio-ecological framework.

**Methods:** We searched PubMed, Embase, Web of Science, CINAHL, and Scopus for U.S.-based studies published between 2010 and 2025 reporting one-year mortality after hip fracture. Studies were included if they evaluated predictors of mortality across pre-injury, perioperative, or post-discharge phases. Data were extracted on study design, population characteristics, mortality outcomes, and risk factors. Predictors examined in ≥3 studies were pooled using random-effects meta-analysis, and narrative synthesis was conducted for predictors with limited data. Methodological quality was assessed using the Joanna Briggs Institute checklist.

**Results:** Twenty-eight studies (n = 835,226) met inclusion criteria. Pooled one-year mortality was 21.8%, ranging from 7.1% to 54.4%. Advancing age and male sex were consistent non-modifiable risk factors. Comorbidity burden, including congestive heart failure, chronic kidney disease, myocardial infarction, and dementia, and measures of frailty and functional impairment were among the strongest predictors, often doubling mortality odds. Perioperative factors such as higher injury severity and delayed surgery, and post-discharge factors including hospital readmission, missed follow-up visits, and postoperative complications, were also associated with increased mortality.

**Conclusion:** One-year hip fracture-related mortality remains high and stems from multifactorial causes. A multi-level, systems-oriented approach may be necessary to meaningfully reduce long-term mortality in this growing and vulnerable population.

## Introduction

Hip fracture remains one of the most serious and life-altering injuries affecting older adults in the United States.^1-7^ Despite substantial progress in orthopedic surgery and geriatric care, the one-year mortality rate following a hip fracture continues to hover between 20% and 30%, making it one of the leading causes of injury-related deaths in this population.^8^ This high fatality rate reflects not only the physiological vulnerability of aging adults but also the cascading medical, functional, and psychosocial consequences that follow a fracture. Survivors frequently experience loss of independence, mobility decline, and an increased likelihood of institutionalization, all of which contribute to poorer long-term outcomes.^9,10^ In addition to the personal suffering endured by patients and families, hip fractures impose a profound financial burden on healthcare systems, with billions spent annually on acute care, rehabilitation, and long-term support services.

Mortality following hip fracture is shaped by complex interactions occurring across three critical temporal phases: pre-injury, peri-operative, and recovery. Each phase captures unique physiological and contextual vulnerabilities that collectively influence survival outcomes. The pre-injury phase reflects the patient’s baseline health status, functional capacity, comorbidities, medication use, nutritional state, and living environment prior to the fracture event.^11^ These factors determine not only the likelihood of sustaining a hip fracture but also the physiological reserve necessary for recovery. The perioperative phase encompasses the period of acute care, from hospital admission to surgical repair, during which the timeliness of surgery, anesthesia type, perioperative complications, and hospital quality of care critically affect survival.^12^ The recovery phase represents the period from hospital discharge to return to community living. Recovery trajectories may be linear (e.g., hospital to home) or non-linear, involving transitions through inpatient rehabilitation or skilled nursing facilities before community reintegration. Long-term survival following hip fracture is shaped during this phase by the continuity of post-discharge care, the availability of social support, the restoration of physical and functional capacity, and the prevention of secondary complications, such as recurrent falls and infections.^13^ A holistic understanding of mortality risk, therefore, requires an integrative approach that bridges these temporal domains rather than viewing them as isolated stages of care.

The socio-ecological model offers a comprehensive perspective on the multifaceted determinants of mortality after hip fracture.^14^ Grounded in public health and behavioral sciences, the socioecological model conceptualizes health outcomes as products of interactions among individual, interpersonal, organizational, community, and policy factors.^15^ In the context of hip fracture recovery, the socio-ecological approach enables a structured exploration of how personal health characteristics intersect with environmental and systemic influences to shape survival trajectories. The socio-ecological model encourages identifying cross-cutting themes— such as health equity, continuity of care, and policy-level factors—that influence outcomes across all levels. By adopting a socio-ecological lens to evaluate potential predictors of one-year mortality in the preoperative, perioperative, and post-discharge phases, a more holistic knowledge of how interventions should be targeted can be obtained.

The aim of this systematic and meta-analytical review is to synthesize available evidence on predictors of one-year mortality following hip fracture among older adults, guided by a socio-ecological framework. This review addresses the question: among U.S. older adults with hip fracture, which patient, interpersonal, health system–level, community, and policy characteristics are associated with one-year mortality in the preoperative, perioperative, and post-discharge phases? Knowledge of these predictors will inform interventions aimed at improving outcomes for older adults after hip fracture.

## Methods

### Search Strategy and Information Sources

This systematic review and meta-analysis were conducted in accordance with the Preferred Reporting Items for Systematic Reviews and Meta-Analyses (PRISMA) guidelines.^16^ We registered the protocol on PROSPERO. We conducted a comprehensive literature search across five electronic databases: PubMed, Embase, Web of Science (WOS), Cumulative Index to Nursing and Allied Health Literature (CINAHL), and Scopus to identify relevant studies published between January 2010 and December 2025. The search strategy was designed to capture studies examining one-year mortality following hip fracture in older adults, with particular emphasis on post-discharge predictors and risk factors. Search terms were tailored to each database’s indexing system while maintaining conceptual consistency across platforms (Table 1). The core search structure combined five key concept domains: (1) hip fracture, (2) older adult, and (3) mortality. In addition to database searches, we manually reviewed reference lists from relevant systematic reviews and key publications to identify additional studies not captured by electronic searches.

**Table 1.** Search Criteria.

|  |  |
| --- | --- |
| PubMed | ("hip fracture"[MeSH] OR "hip fracture")<br>AND ("older adult*" OR elderly OR geriatric)<br>AND ("mortality" OR "survival" OR "death")<br>AND ("1-year" OR "12-month") |
| EMBase | ('hip fracture'/exp OR 'hip fracture')<br>AND ('elderly'/exp OR 'older adult*' OR geriatric)<br>AND ('mortality'/exp OR mortality OR 'death') |
| CINAHL | ("hip fracture")<br>AND ("older adults" OR elderly OR geriatric)<br>AND ("mortality" OR "death") |
| SCOPUS | TITLE-ABS-KEY("hip fracture")<br>AND TITLE-ABS-KEY("older adults" OR elderly OR geriatric)<br>AND TITLE-ABS-KEY("mortality" OR "death") |
| WOS | ("hip fracture")<br>AND ("older adults" OR elderly OR geriatric)<br>AND ("mortality" OR "death") |

### Eligibility Criteria

We included studies that met the following criteria: (1) identified older adults who sustained a hip fracture; (2) used one-year mortality as a primary or secondary outcome, and (3) examined predictors of one-year mortality. Additionally, the studies must focus on the United States population, be published in English, and have full text available. We restricted our review to US-based studies to ensure consistency in healthcare delivery systems, reimbursement structures, and sociodemographic contexts that may influence mortality outcomes. We excluded studies that focused on short-term mortality or long-term mortality without a specific time frame. Additionally, we excluded abstract-only publications, protocols, editorials, opinion pieces, case reports, or case series. Paper published before 2010 were also excluded.

### Study Selection and Data Extraction

The initial database searches yielded 3,860 records across all five databases. After removing 694 duplicates, 3,166 unique records remained for title and abstract screening. Two independent reviewers screened all titles and abstracts using the predefined eligibility criteria. During this phase, 2,925 records were excluded for the following reasons: not US-based (n = 2,675), no mortality outcome (n = 211), and not related to hip fracture (n = 39). Of the remaining 241 articles, 152 studies had either no full text, were not in English, were study protocols, or were case reports/series. We screened the full texts of the remaining 89 articles and excluded 61 papers because they reported 3-month, 6-month, or time-to-death outcomes without information on 1-year mortality. The study selection process is illustrated in a PRISMA flow diagram (Figure 1).

**Figure 1.**
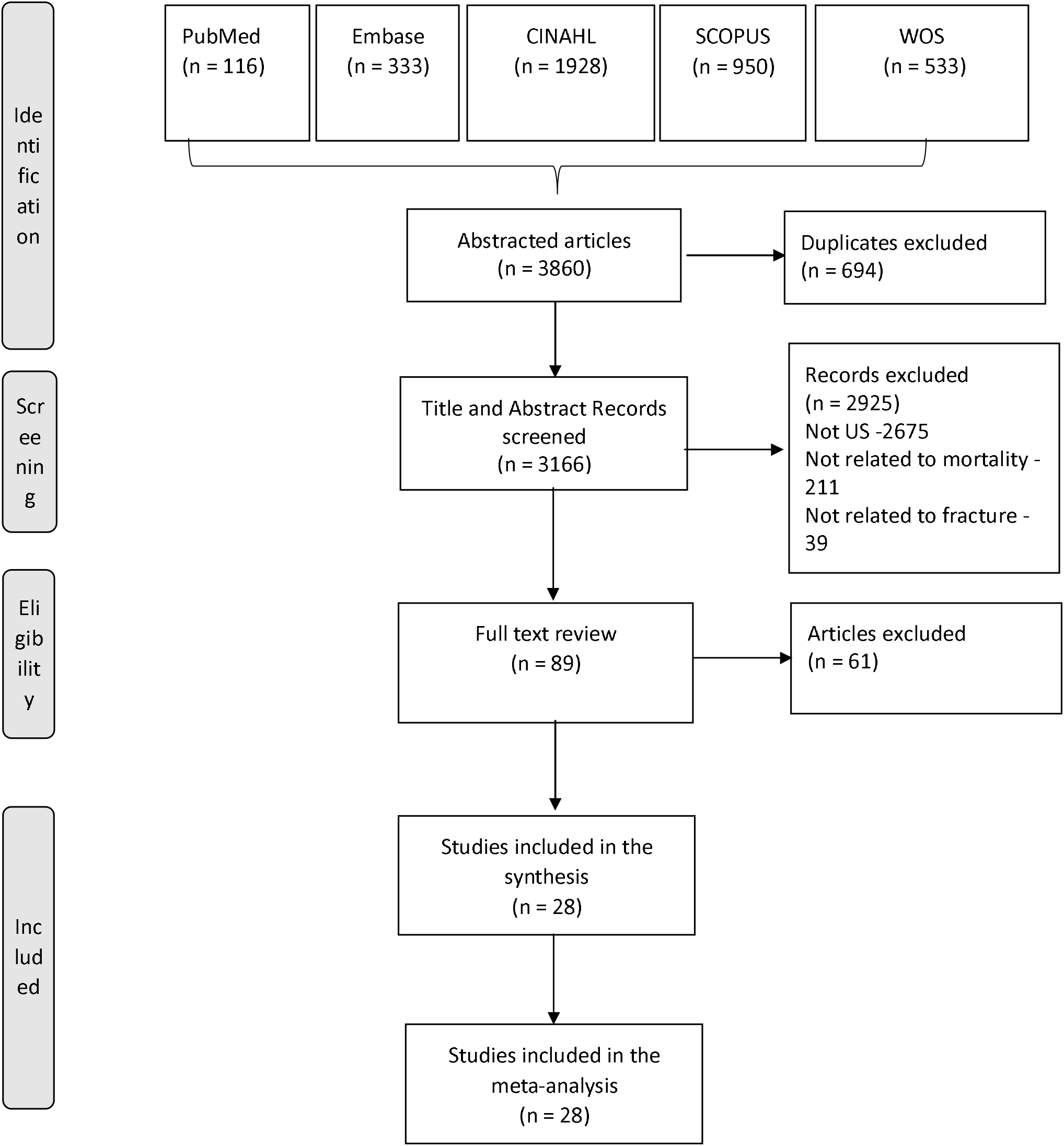
PRISMA diagram showing steps in study selection

**Figure 2.**
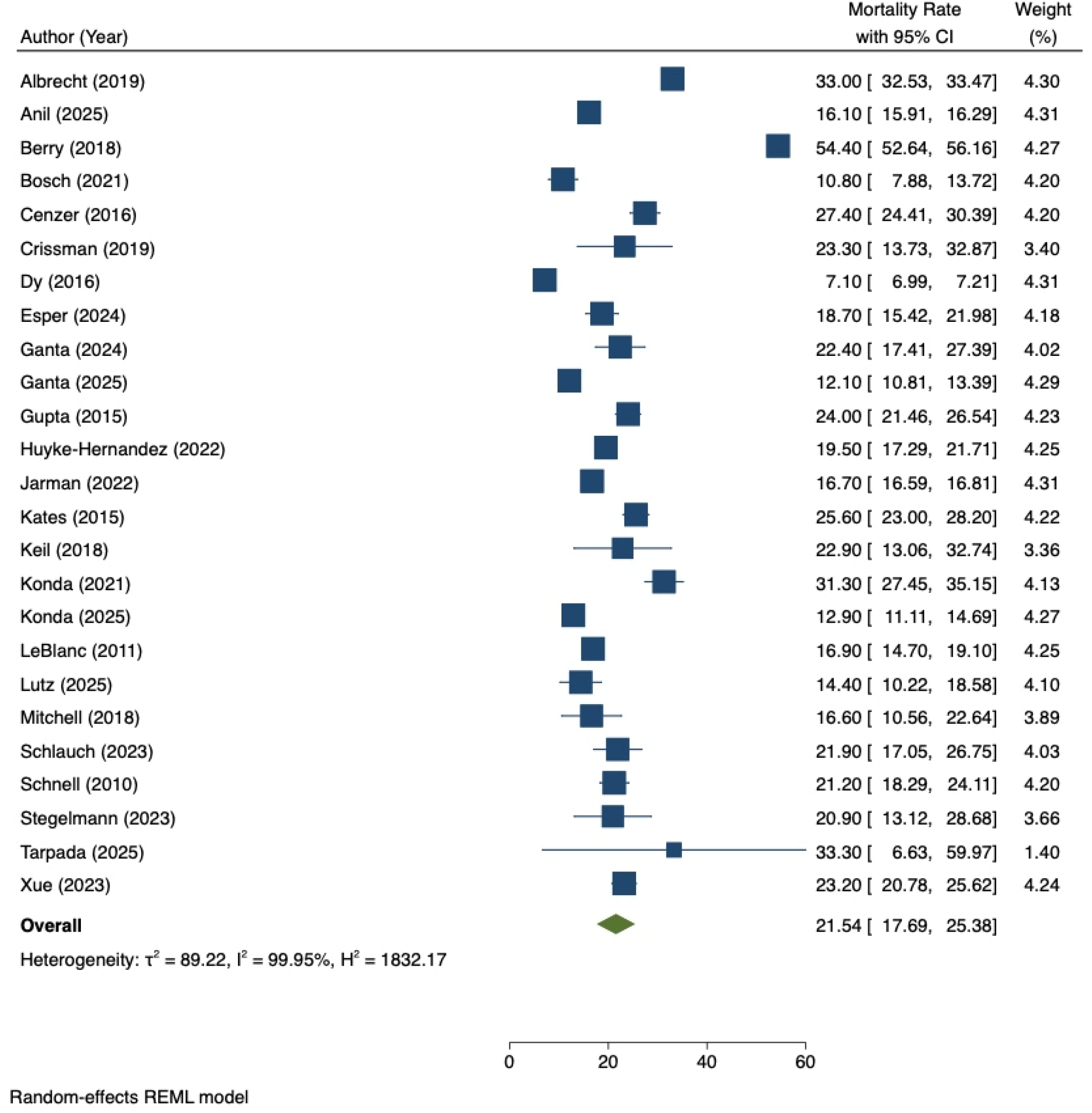
Pooled one-year mortality rate among older adults with hip fractures

### Conceptual Framework and Categorization

All extracted predictors were systematically categorized according to two complementary frameworks: (1) temporal phase (pre-injury, peri-operative, or post-discharge/recovery), and (2) socio-ecological level (individual, interpersonal, organizational, community, or policy). This dual classification system enabled us to identify not only when specific risk factors influence mortality but also at what level of the socio-ecological model these predictors operate.

### Quality Assessment

The methodological quality of included studies was evaluated using the Joanna Briggs Institute (JBI) Critical Appraisal Checklist for Cohort Studies.^17^ This standardized tool assesses the risk of bias across key domains, including the similarity and recruitment of comparison groups, the validity and reliability of exposure and outcome measurement, the identification and management of confounding factors, the completeness and duration of follow-up, and the appropriateness of statistical analysis. The checklist consists of 11 items, each rated as “yes,” “no,” “unclear,” or “not applicable.” For scoring purposes, each “yes” response was assigned one point, yielding a maximum possible score of 11, with higher scores indicating better methodological quality and lower risk of bias. Two independent reviewers conducted all quality assessments, with disagreements resolved by consensus.

### Statistical Analysis and Meta-Analysis

For predictors examined in fewer than three studies or with incompatible effect-size metrics, we conducted a narrative synthesis organized by temporal phase and socio-ecological level. This qualitative synthesis identified patterns, consistencies, and gaps in the evidence base, while highlighting predictors that warrant further investigation. For predictors reported in three or more studies with compatible effect size metrics, we conducted random-effects meta-analyses using the DerSimonian and Laird method.^18^ This approach accounts for both within-study sampling variability and between-study heterogeneity, producing pooled estimates that reflect the expected average effect across diverse study contexts. We calculated pooled odds ratios (ORs), hazard ratios (HRs), or relative risks (RRs) with 95% confidence intervals for each predictor. When studies reported different effect-size metrics for the same predictor, we converted estimates to a common metric when methodologically appropriate or analyzed them separately by metric type.

Heterogeneity across studies was quantified using the I^2^ statistic and Cochran’s Q test.^19,20^ I^2^ values of 25%, 50%, and 75% were interpreted as low, moderate, and high heterogeneity, respectively. ^19,20^ When substantial heterogeneity (I^2^ > 50%) was detected, we conducted sensitivity analyses and, where feasible, subgroup analyses to explore potential sources of variation. ^19,20^ Publication bias was assessed through multiple complementary approaches. Visual inspection of funnel plots was conducted to assess the symmetry of effect-size distributions around the pooled estimates.^21^ Egger’s regression test provided a formal statistical assessment of funnel plot asymmetry, with p < 0.05 indicating potential publication bias. ^21^Additionally, we employed the trim-and-fill method to estimate the number of potentially missing studies due to publication bias and to generate adjusted pooled effect sizes accounting for these hypothetical studies.^22^ All statistical analyses were performed using Stata version 16.^23^ Statistical significance was set at α = 0.05 for all tests. Our reporting adhered to the Meta-analysis of Observational Studies in Epidemiology (MOOSE) guidelines.^24^

## Results

### Study Characteristics

Across the 28 studies, 17 studies used an age benchmark of 65 years and older. The majority of studies (n = 18) employed a retrospective cohort design, while the remaining 10 papers used a matched cohort (n = 5) or a prospective cohort (n = 5). Data sources were mostly institutional (n = 19), while the remaining 9 data sources included national administrative and registry databases, such as the Centers for Medicare & Medicaid Services (CMS) Chronic Conditions Warehouse (CCW), CMS Minimum Data Set (MDS), and the Health and Retirement Study (HRS). Study durations ranged from approximately 2 to 18 years, and sample sizes varied widely, from 12 to 433,169 participants, for a combined total of 835,226 individuals across all 28 studies. Overall study quality was high, with half of the studies receiving the maximum score of 11.

### One-Year Mortality Rates and Variation

Across the 28 included studies, the one-year mortality rates following hip fracture varied substantially, ranging from 7.1% to 54.4% (Table 2). The lowest mortality was observed in a large prospective cohort using SPARCS data (7.1%), while the highest was reported among nursing home residents with advanced dementia (54.4%). Most studies reported one-year mortality rates between approximately 15% and 30%, particularly among community-dwelling older adults aged 65 years and older. Higher mortality estimates were generally observed in cohorts with advanced age (e.g., ≥80 years), significant comorbid conditions, dementia, or institutionalized populations. Using a random effect model, the pooled one-year mortality rate after hip fracture was 21.8%.

**Table 2.**
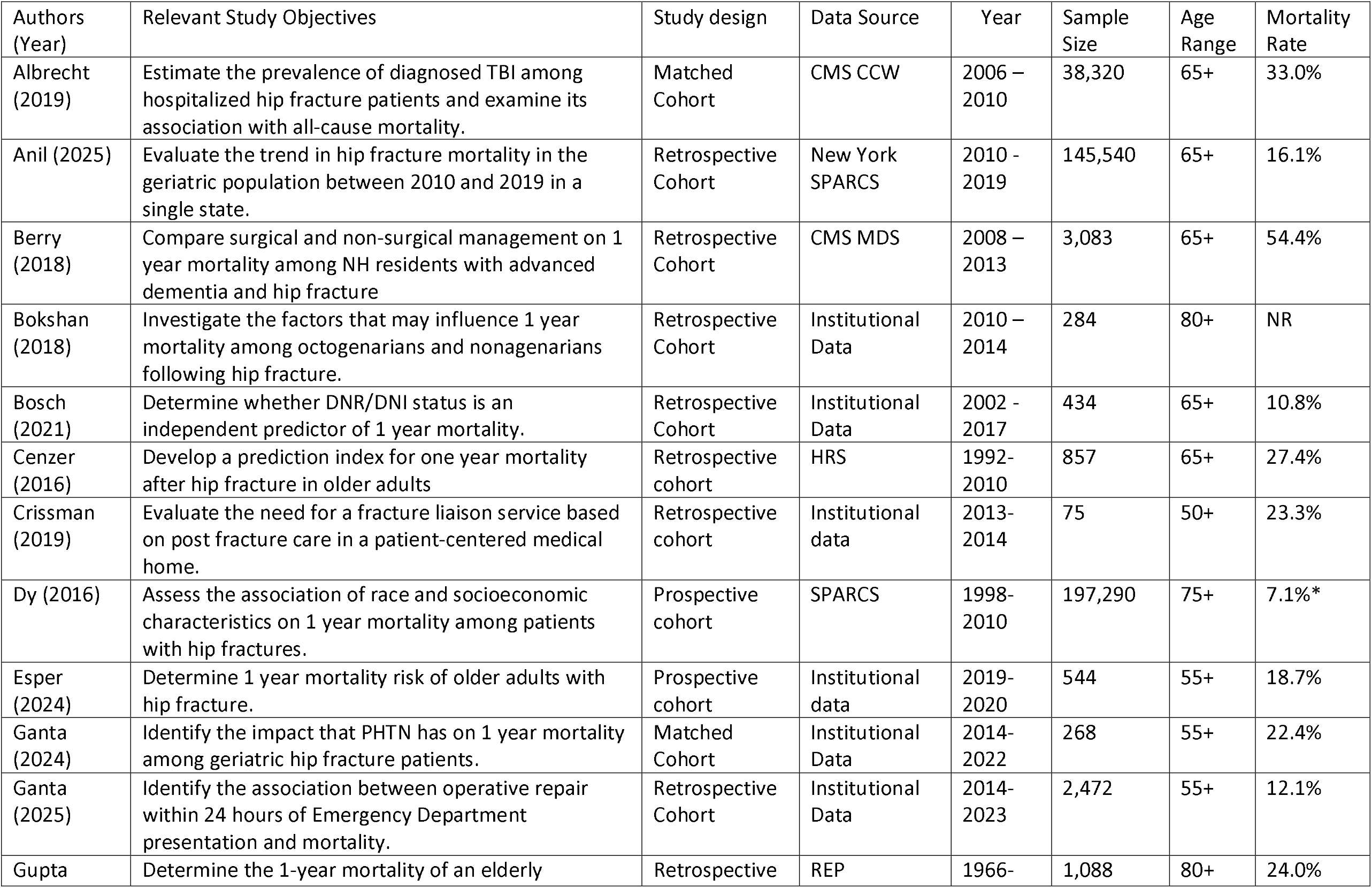
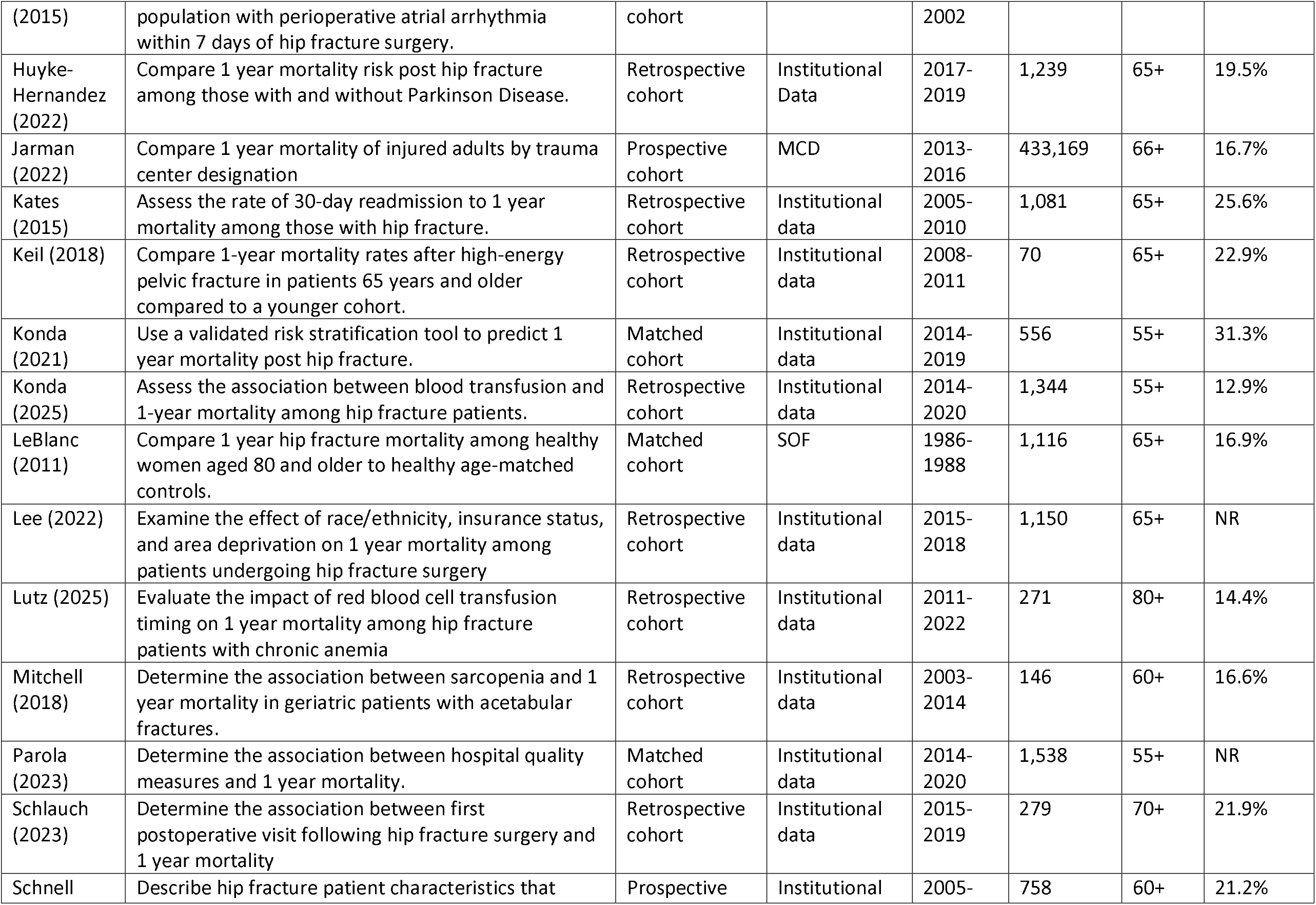
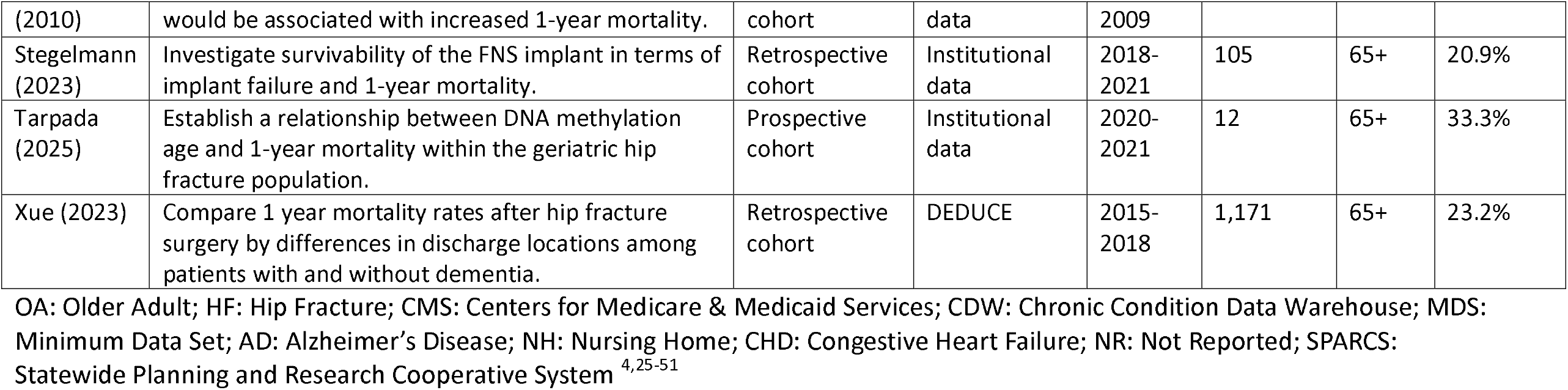
Characteristics of the included studies.

**Table 3.** Synthesis of Pre-Injury Non-modifiable injury factors and their association with one-year hip fracture-related mortality.

| Factor | Author (Year) | Odds or Risk Ratio | Variable Definition | Pattern of Association |
| --- | --- | --- | --- | --- |
| Age | Albrecht (2019) | 1.04 (1.04, 1.04) | Continuous | **Mostly consistent:**<br>Seven of 9 papers reported advancing age is associated with **increased one year mortality risk** |
|  | Keil (2018) | 0.95 (0.85, 1.06) | Continuous |  |
|  | Mitchell (2018) | 1.15 (1.06, 1.25) | Continuous |  |
|  | Schnell (2010) | 1.03 (1.00, 1.06) | Continuous |  |
|  | Xue (2023) | 1.03 (1.01, 1.06) | Continuous |  |
|  | Bosch (2021) | 2.06 (0.93, 4.98) | Category: 85-100 vs 65-84 |  |
|  | Anil (2025) | 1.49 (1.46, 1.51) | Age per decade |  |
|  | Gupta (2015) | 1.40 (1.20, 1.70) | Age per decade |  |
|  | Stegelmann (2023) | 3.71 (1.79, 8.59) | Age per decade |  |
| Race/Ethnicity | Anil (2025) | 1.05 (0.96, 1.15) | Black vs Asian | **Mostly inconsistent:**<br>In the four papers, no race or ethnicity showed elevated one-year mortality risk |
|  | Anil (2025) | 0.93 (0.86, 1.02) | Hispanic vs Asian |  |
|  | Anil (2025) | 0.92 (0.69, 1.21) | Nat.Am vs Asian |  |
|  | Anil (2025) | 0.82 (0.76, 0.89) | White vs Asian |  |
|  | Lee (2022) | 1.81 (0.64, 5.09) | NHB vs Other Races |  |
|  | Lee (2022) | 0.77 (0.49, 1.20) | NHW vs NHB |  |
|  | Lee (2022) | 1.39 (0.54, 3.59) | NHW vs Other Races |  |
|  | Parola (2023) | 2.00 (1.20, 2.86) | White vs Non-White |  |
|  | Xue (2023) | 0.99 (0.69, 1.39) | White vs Non-White |  |
| Sex | Anil (2025) | 1.42 (1.39, 1.45) | Male vs Female | **Mostly consistent:**<br>Five of the 9 papers reported **increased one-year mortality risk** among male older adults |
|  | Bosch (2021) | 1.45 (0.76, 2.92) | Male vs Female |  |
|  | Cenzer (2016) | 1.97 (1.41, 2.75) | Male vs Female |  |
|  | Gupta (2015) | 2.20 (1.60, 2.90) | Male vs Female |  |
|  | Mitchell (2018) | 0.56 (0.15, 2.12) | Male vs Female |  |
|  | Parola (2023) | 1.4 (0.99, 2.00) | Male vs Female |  |
|  | Schnell (2010) | 1.56 (1.02, 2.40) | Male vs Female |  |
|  | Stegelmann (2023) | 0.49 (0.18, 1.41) | Male vs Female |  |
|  | Xue (2023) | 1.85 (1.35, 2.56) | Male vs Female |  |

Nat.Am: Native American; NHB: Non-Hispanic Black; NGW: Non-Hispanic White

### Pre-Injury Non-Modifiable Individual Factors

Advancing age was the most consistently identified non-modifiable risk factor, with 7 of 9 studies demonstrating a significant association between increasing age and higher one-year mortality, regardless of whether age was modeled continuously or categorically (Table 4). The meta-analysis of non-modifiable individual factors, using the four studies that treated age as a continuous variable, showed that for every additional year, the odds of mortality increased by 6%. (Pooled OR: 1.06; 95% CI: 1.01 – 1.11; Figure 3). For every 10-year interval, the mortality odds increase by 91% (Pooled OR: 1.91; 95% CI: 1.07 – 3.42). Male sex was also commonly associated with increased mortality risk, with 5 of 9 studies reporting significantly higher odds among men compared to women. The pooled odds of mortality by sex showed that males had 38% increased mortality odds compared to females (Pooled OR: 1.38; 95% CI: 1.03 – 1.85). In contrast, race and ethnicity demonstrated inconsistent findings across studies, with most analyses showing no statistically significant differences in one-year mortality across racial or ethnic groups.

**Table 4.** Synthesis of Pre-Injury modifiable injury factors and their association with one-year hip fracture-related mortality.

| Factor | Author (Year) | Odds or Risk Ratio | Variable Definition | Pattern of Association |
| --- | --- | --- | --- | --- |
| Chronic comorbidities | Anil (2025) | 1.07 (1.07, 1.07) | Elixhauser (Continuous) | **Mostly consistent:**<br>Seven of 9 papers reported **increasing one-year mortality risk** with increasing number of comorbid conditions. |
|  | Bokshan (2018) | 5.38 (1.70, 16.60)<br><sup>a</sup> 2.06 (1.03, 4.10)<br><sup>b</sup> | CCI 3+ vs 0 |  |
|  | Bosch (2021) | 1.21 (0.62, 2.41) | CCI 6+ vs <6 |  |
|  | Ganta (2024) | 1.39 (1.14, 1.69) | CCI (Continuous) |  |
|  | Gupta (2015) | 1.90 (1.10, 3.30) | ASA3-5 vs 1-2 |  |
|  | Keil (2018) | 1.03 (1.00, 1.07) | CCI (Continuous) |  |
|  | Mitchell (2018) | 1.09 (0.73, 1.62) | CCI (Continuous) |  |
|  | Schnell (2010) | 2.19 (1.32, 3.64)<br>1.36 (0.81, 2.28) | CCI 4+ vs 0-1<br>CCI2-3 vs 0-1 |  |
|  | Xue (2023) | 1.09 (1.04, 1.15) | CCI (Continuous) |  |
| Specific Chronic Medical Conditions |  |  |  |  |
| Hypertension | Gupta (2015) | 0.80 (0.60, 1.00) | Hypertension | **Consistent:**<br>No association |
|  | Albrecht (2019) | 1.00 (0.95, 1.06) | Hypertension |  |
| Chronic Kidney Disease | Cenzer (2016) | 2.54 (1.67, 3.86) | CKD | **Consistent:**<br>Strong association |
|  | Gupta (2015) | 2.00 (1.50, 2.80) | CKD |  |
| MI | Albrecht (2019) | 1.19 (1.15, 1.23) | Ischemic HD | **Consistent:**<br>Strong association |
|  | Cenzer (2016) | 1.55 (1.01, 2.39) | myocardial infarction |  |
|  | Gupta (2015) | 1.90 (1.50, 2.50) | myocardial infarction |  |
| DM | Gupta (2015) | 1.40 (1.00, 2.00) | Diabetes | **Inconsistent:**<br>1 of 2 papers reported an association |
|  | Cenzer (2016) | 1.34 (0.96, 1.87) | Diabetes |  |
| Congestive Heart Failure | Cenzer (2016) | 2.95 (2.15, 4.05) | Binary | **Consistent:**<br>Strong association<br>Increased one-year mortality risk |
|  | Gupta (2015) | 1.70 (1.30, 2.20) |  |  |
| CVA | Cenzer (2016) | 1.32 (0.94, 1.85) | CVA | **Consistent:**<br>No association |
|  | Gupta (2015) | 1.20 (0.90, 1.60) | CVA |  |
|  | Albrecht (2019) | 1.01 (0.98, 1.05) | CVA |  |
| Dementia | Albrecht (2019) | 1.61 (1.56, 1.66) | Binary | **Consistent:**<br>Strong association<br>Increased one- |
|  | Cenzer (2016) | 1.69 (1.14, 2.50) |  |  |
|  | Gupta (2015) | 2.40 (1.80, 3.10) |  |  |
|  | Xue (2023) | 1.78 (1.33, 2.38) |  | year mortality risk |
| Obesity | Anil (2025) | 1.11 (1.04, 1.18) | Obesity | **Mostly consistent:**<br>2 of 3 paper reported no association |
|  | Gupta (2015) | 0.80 (0.50, 1.30)<br>0.90 (0.70, 1.30) | Obese vs. Normal<br>Overweight vs. Normal |  |
|  | Stegelmann (2023) | 0.98 (0.90, 1.06) | BMI (continuous) |  |
| Other Chronic Medical Conditions | Albrecht (2019) | 1.28 (1.24, 1.33) | Atrial Fibrillation | **Mostly consistent:**<br>6 of 11 listed comorbidity are independently associated with increased one-year mortality risk |
|  | Albrecht (2019) | 1.57 (1.49, 1.66) | Coagulation disorder |  |
|  | Albrecht (2019) | 1.08 (1.05, 1.12) | Depression |  |
|  | Cenzer (2016) | 1.44 (0.97, 2.15) | Any malignancy |  |
|  | Cenzer (2016) | 1.42 (1.04, 1.94) | COPD |  |
|  | Cenzer (2016) | 1.78 (1.22, 2.59) | PVD |  |
|  | Cenzer (2016) | 1.39 (0.76, 2.55) | Ulcers |  |
|  | Ganta (2024) | 2.72 (1.24, 7.55) | Pulmonary HTN |  |
|  | Gupta (2015) | 1.00 (0.70, 1.40) | Atrial Arrhythmia |  |
|  | Gupta (2015) | 1.20 (0.50, 3.30) | Obstructive Sleep Apnea |  |
|  | H-Hernandez (2022) | 1.52 (0.79, 2.91) | Parkinson |  |
| Do Not Resuscitate or Intubate order | Bosch (2021) | 9.69 (4.02, 27.8) | DNR/DNI vs full code | **Insufficient data:**<br>Data from a single paper |
| Alcohol | Xue (2023) | 0.81 (0.53, 1.24) | Current alcohol y/n | **Insufficient data:**<br>1 paper; No association |
| Smoking | Anil (2025) | 1.04 (1.01, 1.07) | Current smoker: Yes vs. No | **Mostly consistent:**<br>2 of 3 paper reported no association |
|  | Mitchell (2018) | 0.50 (0.16, 1.64) |  |  |
|  | Xue (2023) | 1.45 (0.85, 2.47) |  |  |
| Frailty | Gupta (2015) | 1.60 (1.10, 2.20) | Underweight vs Normal Weight | **Consistent:**<br>Strong association<br>Increased one-year mortality risk |
|  | Mitchell (2018) * | 0.02 (0.00, 0.88) | Psoas:Lumbar Vertebral Index |  |
|  | Mitchell (2018) | 4.04 (1.62, 10.10) | Sarcopenia |  |
| Functional Limitations | Schnell (2010) | 1.84 (0.99, 3.44)<br>1.60 (0.93, 2.76) | ADL<br>Dep. vs Independent<br>Partial vs Independent | **Mostly Consistent:**<br>2 of 3 paper reported that having difficulty or being dependent is |
|  | Schnell (2010) | 2.79 (1.24, 6.27)<br>2.17 (0.99, 4.42) | Parker Mobility Score<br>Low vs High<br>Medium vs high |  |
|  | Cenzer (2016) ** | 1.82 (1.56 – 2.22) | Difficulty vs. No Difficulty | associated with one-year mortality risk. |

a: Nonagenarian; b: Octogenarians; COPD: Chronic Obstructive Pulmonary Disease; PVD: Peripheral vascular Disease; \*Higher scores indicate more reserve and less frailty. \*\*Odds ratios reported are pooled estimates from computed crude odds ratios using counts and percents in the publication.

### Pre-Injury Modifiable Individual Factors

Among modifiable pre-injury factors, increasing comorbidity burden, measured using indices such as the Charlson Comorbidity Index (CCI), Elixhauser score, or ASA classification, was consistently associated with higher one-year mortality in 7 of 9 studies (Table 5). Using three of the studies that assessed the relationship of the Charlson comorbidity index (as a continuous variable) and one-year mortality, a unit increase in the Charlson comorbidity index score was associated with 19% increased odds of mortality (Pooled OR: 1.19; 95% CI: 1.01 – 1.40; Figure 4). Also, several specific chronic conditions, including chronic kidney disease, myocardial infarction, congestive heart failure, and dementia, demonstrated strong and consistent associations with increased mortality risk. Specifically, the congestive heart failure (Pooled OR: 2.19; 95% CI: 1.60 – 2.99) and dementia (Pooled OR: 1,84; 95% CI: 1.54 – 2.21) both have approximately two-fold odds of mortality (Figure 4). Measures of frailty, including sarcopenia, albumin, and underweight, were consistently associated with mortality across studies, with the pooled OR showing a two-fold mortality odds (Pooled OR: 2.16; 95% CI: 1.30 – 3.58; Figure 5). In contrast, hypertension, cerebrovascular accident, obesity, alcohol use, and smoking generally showed no consistent association. Functional limitations were mostly associated with increased mortality risk across the studies, with the pooled OR showing a two-fold mortality odds (Pooled OR: 1.97; 95% CI: 1.61 – 2.42).

**Table 5.** Synthesis of Pre-Injury factors in the interpersonal, organizational, community, and policy domains of the socio-ecological model.

| Factor | Author (Year) | Odds or Risk Ratio | Variable Definition | Pattern of Association |
| --- | --- | --- | --- | --- |
| **Interpersonal Level** |  |  |  |  |
| Dyadic Support | Cenzer (2016) | 0.92 (0.67, 1.26)<br>2.22 (1.56, 3.16) | Married/Partnered (Yes vs No)<br>Proxy Interview Respondent (Yes vs No) | **Insufficient data:**<br>Data from a single paper |
| **Community Level** |  |  |  |  |
| Residence before Injury | Cenzer (2016) | 2.9 (1.81, 4.64) | Nursing Facility vs Home | **Consistent:**<br>Strong association between nursing home residence and increased one-year mortality risk |
|  | Xue (2023) | 1.12 (0.73, 1.71)<br>0.95 (0.40, 2.25)<br>1.59 (1.06, 2.39) | Hospital vs Home<br>Others vs Home<br>Nursing Facility vs Home |  |
| Environment | Lee (2022) | 1.32 (0.44, 3.94)<br>1.50 (0.84, 2.68) | Area Deprivation Index<br>25-75th vs 0-25<sup>th</sup><br>75-100th vs 0-25<sup>th</sup> | **Insufficient data:**<br>Data from a single paper |
| **Policy Level** |  |  |  |  |
| Insurance | Lee (2022) | 1.69 (1.10, 2.63)<br>0.76 (0.28, 2.13) | Medicare vs MA<br>Medicare vs. Others | **Inconsistent:**<br>Medicare comparison with other insurance not significant |
|  | Xue (2023) | 1.32 (0.90, 1.94) | Medicare vs Others |  |

MA: Medicaid Advantage

**Table 5.** Synthesis of perioperative and post-discharge factors in the interpersonal, organizational, community, and policy domains of the socio-ecological model.

| Factor | Author (Year) | Odds or Risk Ratio | Variable Definition | Pattern of Association |
| --- | --- | --- | --- | --- |
| Peri-operative Individual |  |  |  |  |
| Fracture Pattern | Mitchell (2018) | 0.91 (0.43, 1.96) | pelvic ring injury | **Consistent:**<br>No association between fracture pattern and one-year mortality risk |
|  | Stegelmann (2023) | 0.92 (0.30, 2.57) | Displaced fracture vs non-displaced |  |
|  | Stegelmann (2023) | 0.76 (0.16, 2.62) | Pauwels III vs I-II |  |
| Injury Severity | Albrecht (2019) | 1.24 (1.14, 1.35) | Coexisting Traumatic Brain Injury | **Consistent:**<br>Strong association between injury severity and one-year mortality risk |
|  | Esper (2024) | 2.11 (1.12, 3.98) | STTGMA High Risk (Yes or No) |  |
|  | Mitchell (2018) | 1.10 (1.05, 1.15) | Injury Severity Score (Continuous) |  |
|  | Parola (2023) | 1.20 (1.17, 1.28) | STTGMA (Continuous) |  |
| Peri-operative Organizational Level |  |  |  |  |
| Time to Surgery | Bokshan (2018) | 10.6 (2.10, 53.3) | 3+ days vs 0-2 days | **Mostly consistent:**<br>2 of 3 paper reported strong association between delay and one-year mortality |
|  | Ganta (2024) | 8.98 (6.05, 13.32) | 3+ days vs 0-2 days |  |
|  | Xue (2023) | 1.33 (0.78, 2.28) | 3+ days vs 0-2 days |  |
| Surgery Type | Xue (2023) | 0.99 (0.75, 1.33)<br>0.13 (0.03, 0.54) | Hemiarthroplasty vs ORIF<br>THA vs ORIF | **Insufficient data:**<br>Data from a single paper |
| Peri-operative Blood Transfusion | Konda (2025) | 1.29 (0.86, 1.93) | Binary | **Inconsistent:**<br>1 of 2 papers reported an association |
|  | Lutz (2025) | 2.93 (1.41, 6.07) | Binary |  |
| Trauma level designation | Jarman (2022) | 0.94 (0.79, 1.14)<br>0.99 (0.83, 1.18)<br>0.96 (0.81, 1.14) | Levels 1 vs 4<br>Levels 2 vs 4<br>Levels 3 vs 4 | **Insufficient data:**<br>Data from a single paper |
| Post-discharge Individual and Organizational Factors |  |  |  |  |
| Post-op AF Complication (Individual) | Gupta (2015) | 2.10 (1.40, 3.10) | Binary | **Consistent:**<br>Strong association between post-operative events and |
| Missed first POV (Individual) | Schlauch (2023) | 7.32 (3.90, 13.73) | Binary |  |
| Re-admission<br>(Organizational) | Kates (2015) | 4.52 (3.09, 6.61) | Binary | increased one-year mortality<br>risk |

POV: Post-Operative Visit; STTGMA: Score for Trauma Triage in Geriatrics and Middle Age

**Figure 4.**
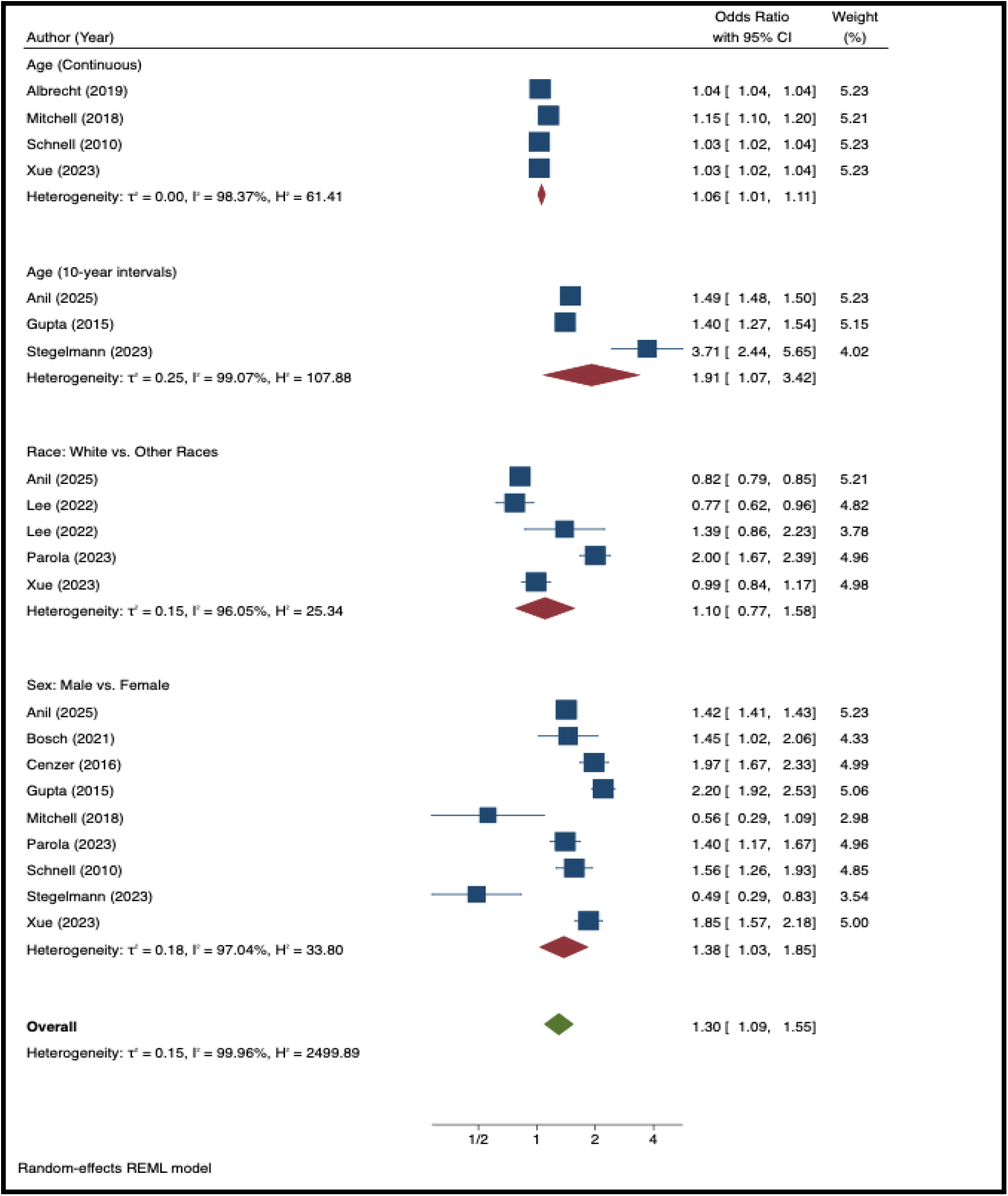
Pooled effect assessing the association of pre-operative demographic characteristics with one-year hip fracture mortality.

**Figure 5.**
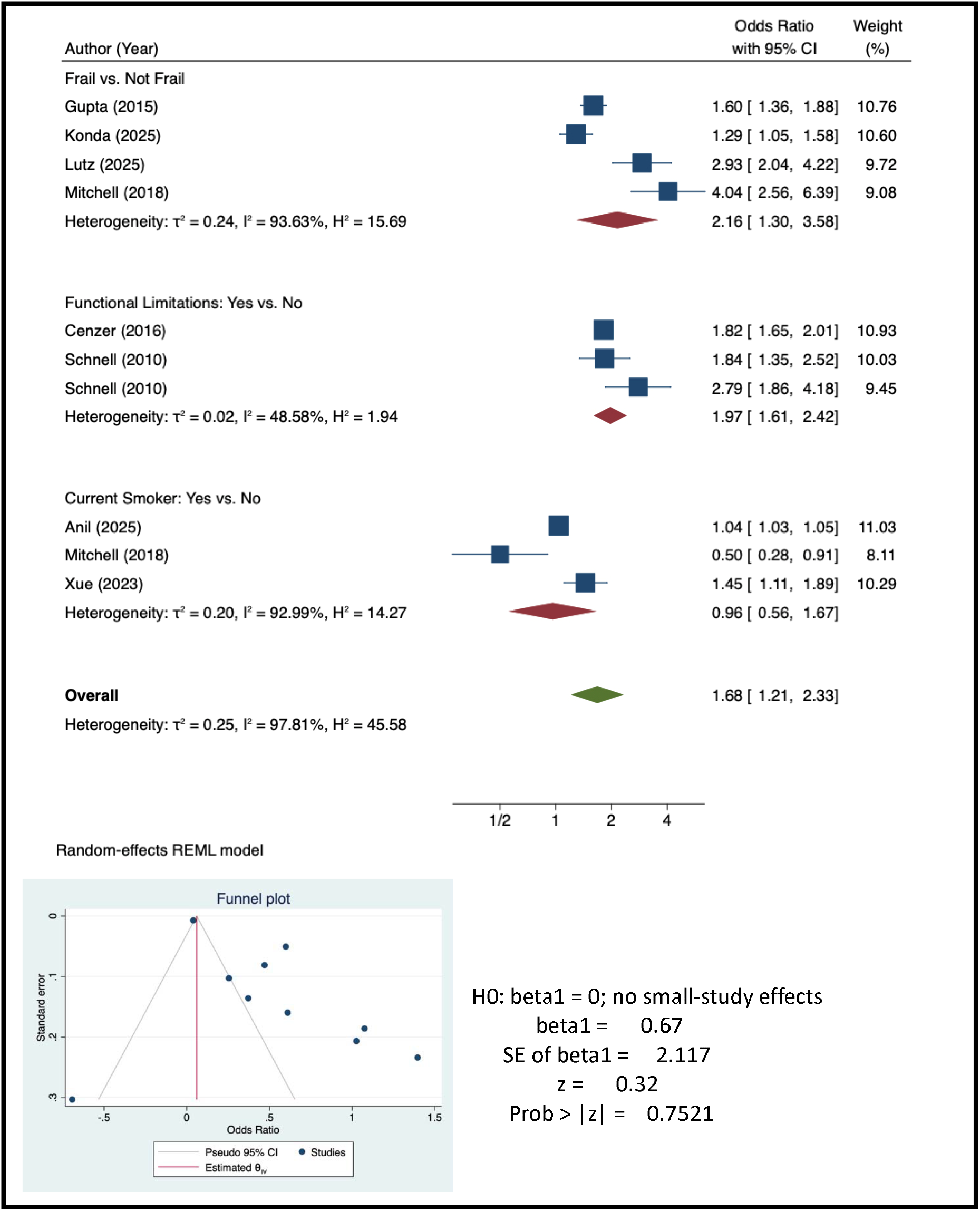
Pooled effect size assessing the association of pre-operative modifiable risk factors with one-year hip fracture mortality.

**Figure 6.**
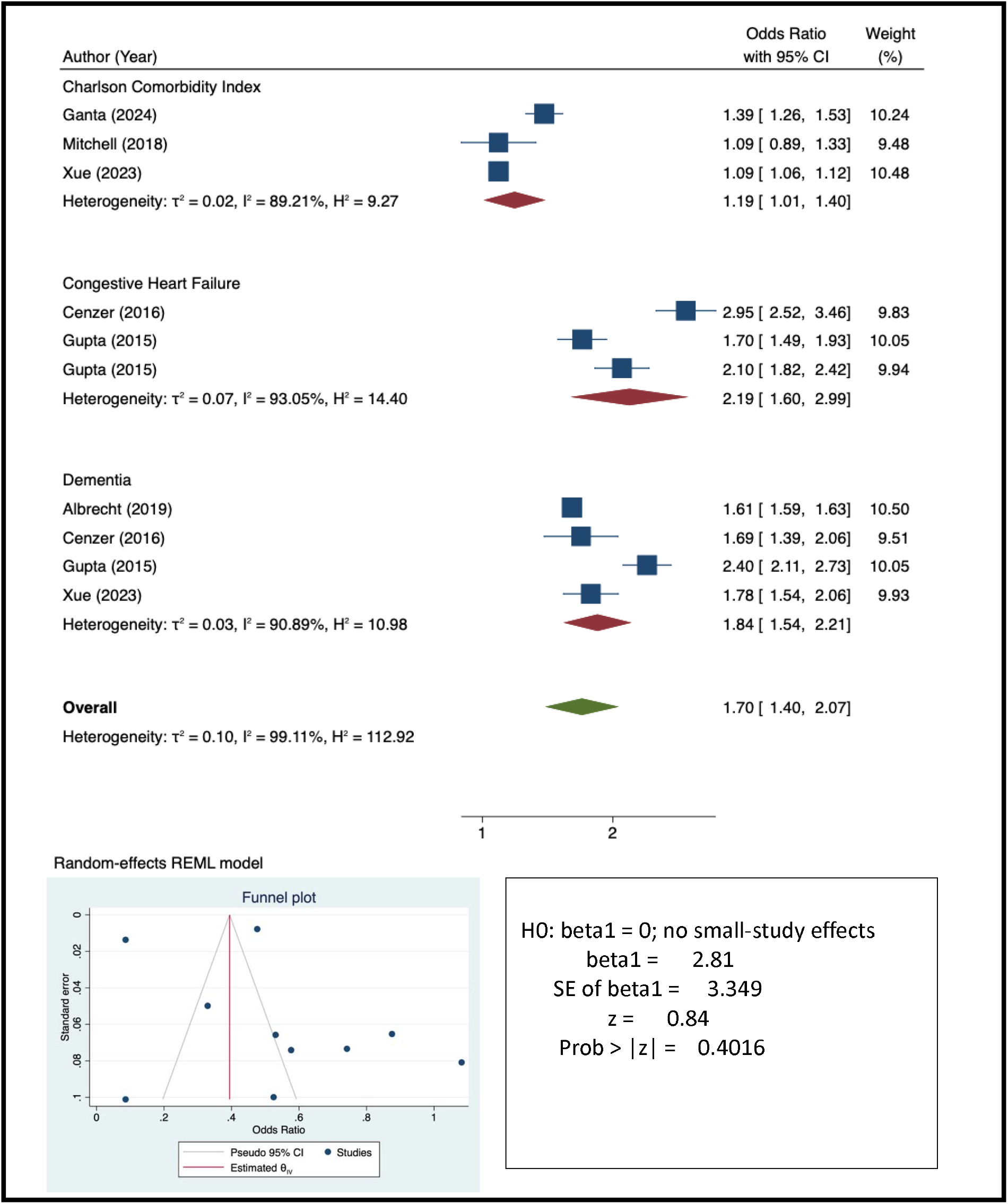
Pooled effect size assessing the association of pre-operative chronic comorbid factors with one-year hip fracture mortality.

**Figure 7.**
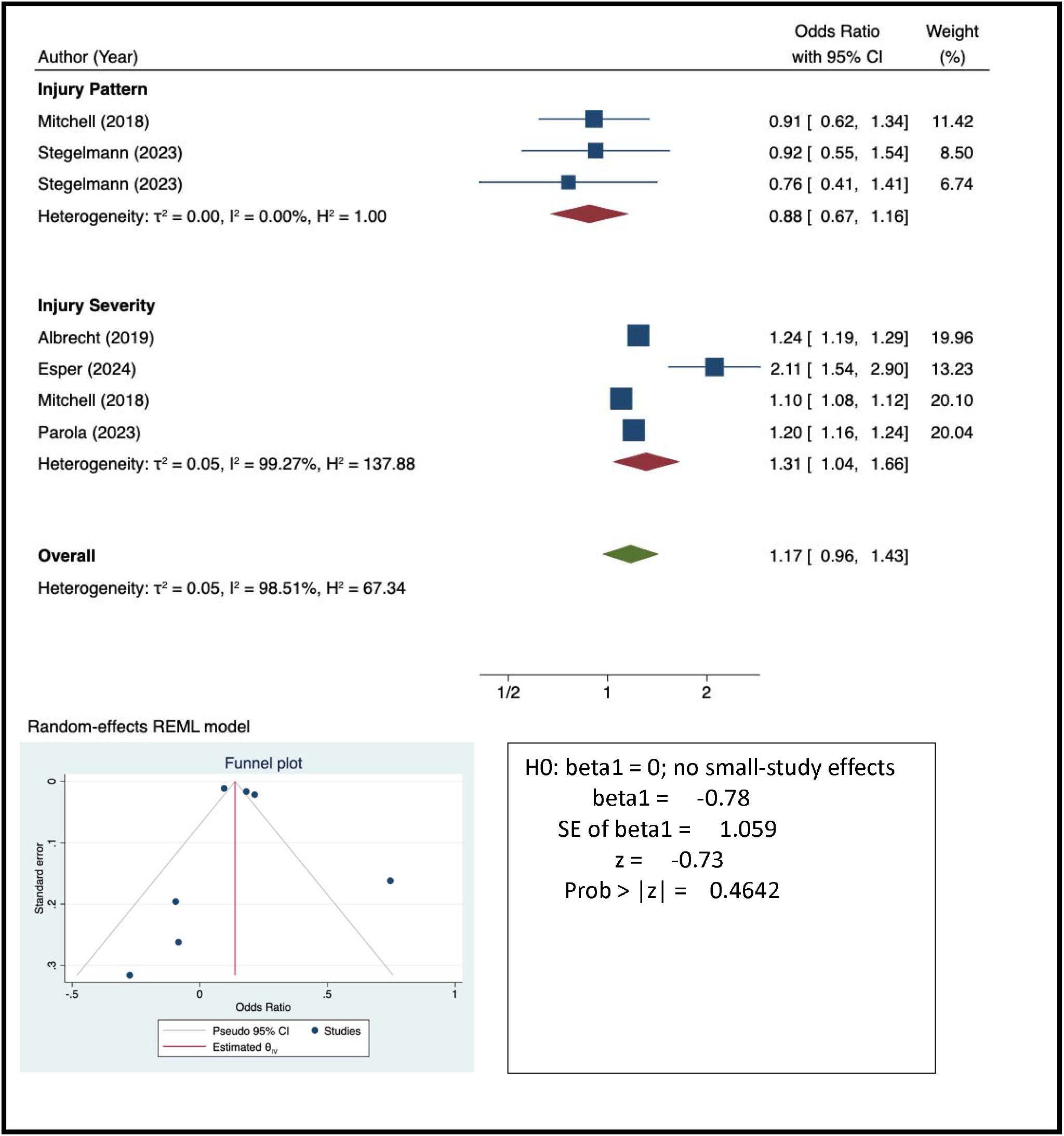
Pooled effect size assessing the association of peri-operative injury pattern and severity with one-year hip fracture mortality.

### Pre-Injury Interpersonal, Community, and Policy Factors

At the interpersonal level, evidence was limited, with dyadic support examined in a single study that yielded mixed findings (Table 5). At the community level, pre-injury residence in a nursing facility was consistently associated with increased one-year mortality compared to residence at home. Environmental factors such as area deprivation index and policy-level factors, including insurance status, demonstrated inconsistent or non-significant associations across the limited studies available.

### Perioperative and Post-Discharge Factors

Fracture pattern was not associated with one-year mortality across studies (Table 6). In contrast, injury severity measures—including coexisting traumatic brain injury, Injury Severity Score, and STTGMA risk classification—were consistently associated with increased mortality. A unit increase in injury severity score was associated with 31% increased odds of mortality (Pooled OR: 1.31; 95% CI: 1.04 – 1.66; Figure 5). Delays of three or more days to surgery were associated with a substantially increased risk of mortality in two of three studies. Evidence regarding the type of surgery, perioperative blood transfusion, and trauma center designation was limited or inconsistent. Post-discharge factors demonstrated strong associations: postoperative atrial fibrillation, missed first postoperative visit, and hospital readmission were all associated with markedly increased one-year mortality.

## Discussion

We set out to assess the predictors of one-year mortality following hip fracture across the pre-injury, perioperative, and post-discharge phases, using the socio-ecological model as a unifying framework. Across 28 studies comprising more than 835,000 individuals, we found that mortality is shaped by determinants operating at multiple levels. At the individual pre-injury level, advancing age, male sex, increasing comorbidity burden, congestive heart failure, dementia, frailty, and functional limitations were consistently associated with increased mortality. Community-level factors, particularly nursing home residence prior to injury, were also strongly associated with worse outcomes. During the perioperative phase, injury severity and surgical delay emerged as important predictors, while fracture pattern itself was not consistently associated with mortality. In the post-discharge phase, postoperative atrial fibrillation, missed follow-up visits, and hospital readmission were strongly linked to increased one-year mortality. Together, these findings suggest that one-year mortality after hip fracture reflects a continuum of vulnerability that spans biological, functional, and system-level domains.

We observed substantial variation in reported one-year mortality rates, ranging from 7.1% to 54.4%, with a pooled estimate of 21.8%. While most community-dwelling cohorts demonstrated mortality rates between 15% and 30%, markedly higher rates were seen among older adults with advanced dementia or those residing in nursing facilities. A recent study further demonstrated significant geographic variability in one-year hip fracture mortality across regions, underscoring the influence of contextual and system-level factors beyond individual risk.^8^ These findings reinforce that hip fracture mortality is not solely a function of age or comorbidity, but rather a multi-level phenomenon shaped by patient vulnerability, injury characteristics, care processes, and broader health system and community contexts.^5,52,53^ The wide variation in mortality rates likely reflects differences in baseline frailty, access to timely surgery, post-acute care resources, and regional healthcare infrastructure. Recognizing mortality as a multi-level outcome, therefore, shifts the focus from isolated predictors to coordinated interventions across the care continuum.

Pre-injury factors emerged as some of the most robust predictors of one-year mortality. Advancing age showed a dose-response relationship, with each additional year increasing the odds of mortality. Also, male sex was independently associated with worse one-year survival, a finding consistently reported in large population-based studies despite women having a substantially higher lifetime risk of osteoporosis and hip fracture.^54,55^ This apparent paradox has been well documented: although women sustain more hip fractures due to lower bone mineral density and postmenopausal bone loss, men experience disproportionately higher mortality after fracture.^54^ Several explanations have been proposed. Men who sustain hip fractures are often older at presentation, have a higher burden of cardiovascular and systemic comorbidities, and may have fractures occurring in the setting of greater underlying frailty or delayed medical engagement.^56,57^ Additionally, hip fracture in men may represent a marker of more advanced physiologic decline, whereas in women it may more commonly reflect osteoporosis-related skeletal fragility alone.^56,57^

Beyond demographic characteristics, comorbidity burden demonstrated a clear graded association with one-year mortality, reinforcing prior evidence that cumulative disease burden meaningfully shapes outcomes after hip fracture.^58,59^ Indices such as the Charlson Comorbidity Index and the Elixhauser index have repeatedly been shown to predict short- and long-term mortality in orthopedic and geriatric populations, reflecting the additive physiologic strain imposed by multi-organ dysfunction.^60-62^ In our synthesis, specific conditions—including congestive heart failure, chronic kidney disease, prior myocardial infarction, and dementia— were particularly influential, likely because they represent systemic vulnerability affecting cardiovascular reserve, metabolic stability, inflammatory response, and cognitive capacity to participate in rehabilitation. Notably, measures of frailty and baseline functional impairment were among the strongest predictors, nearly doubling mortality odds. This aligns with emerging literature demonstrating that frailty—conceptualized as diminished physiologic reserve and impaired stress response—outperforms individual comorbidities in predicting postoperative mortality. ^63-66^

During the perioperative phase, injury severity consistently predicted one-year mortality, whereas fracture pattern did not. Coexisting traumatic brain injury, higher Injury Severity Scores, and elevated STTGMA risk classifications were all associated with increased mortality risk, underscoring the prognostic value of global injury burden rather than fracture morphology alone.^67-69^ Delays of three or more days to surgery were associated with substantially increased mortality in most studies examining this relationship, supporting existing recommendations for timely operative management.^70,71^ The perioperative window, therefore, represents a critical opportunity for risk stratification and optimization, particularly among patients with high injury severity or delayed operative intervention.

The post-discharge factors demonstrated some of the strongest associations with one-year mortality. Postoperative atrial fibrillation, missed first postoperative visits, and hospital readmissions were each associated with markedly increased mortality risk. These findings highlight that survival after hip fracture is not determined solely during the index hospitalization but continues to be shaped by recovery trajectories, complication surveillance, and care continuity. Missed follow-up visits may represent both patient-level vulnerability and system-level gaps in care coordination.^72^ Similarly, readmission likely reflects unresolved medical instability or inadequate post-acute support. These results emphasize that the post-discharge period is a high-risk phase and may represent an underutilized target for intervention. Structured transitional care programs, enhanced follow-up systems, and proactive management of postoperative complications may meaningfully influence longer-term survival.^73^

Despite the breadth of included studies, important gaps remain. Interpersonal, community, and policy-level determinants were underexamined, with limited and inconsistent data on social support, area deprivation, and insurance status. Few studies explored structural inequities, regional healthcare variation, or access to rehabilitation services in depth. Additionally, many analyses relied on single-institution cohorts, limiting generalizability. There was also substantial heterogeneity in variable definitions, adjustment models, and mortality reporting, which constrained pooled analyses. Notably, relatively few studies examined modifiable system-level interventions or evaluated causal pathways linking pre-injury vulnerability to post-discharge outcomes. Future research should prioritize multi-level modeling approaches, standardized definitions, and prospective studies that integrate biological, functional, and social determinants to better understand mechanisms underlying mortality risk.

Our study’s limitations warrant consideration. Clinical and methodological heterogeneity across studies was substantial, including differences in populations, adjustment strategies, and definitions of exposures and outcomes. Most studies were retrospective and observational, limiting causal inference. Some domains, particularly interpersonal and policy-level factors, were informed by a small number of studies. Finally, publication bias and residual confounding cannot be excluded. Despite these limitations, this review has several strengths. We applied a socio-ecological framework to systematically organize predictors across the care continuum and integrated meta-analytic estimates where appropriate. The inclusion of over 835,000 individuals across diverse settings enhances the robustness of our findings. This study represents one of the few studies that used a validated framework to assess the preinjury, preoperative, and post-discharge predictors of one-year mortality.

## Conclusion

One-year mortality following hip fracture remains high, affecting approximately one in five older adults, and is shaped by determinants operating across pre-operative,, perioperative, and post-discharge domains. Age, male sex, comorbidity burden, dementia, frailty, injury severity, surgical delay, and post-discharge complications emerged as the most consistent predictors. Importantly, mortality after hip fracture should be conceptualized not as a single event outcome but as the culmination of multi-level vulnerabilities and care processes across the recovery trajectory. Interventions aimed at reducing mortality must therefore extend beyond the operating room to include comprehensive geriatric assessment, timely surgery, coordinated transitional care, and targeted support for high-risk individuals. A multi-level, systems-oriented approach may be necessary to meaningfully reduce long-term mortality in this growing and vulnerable population.

## Data availability

All data produced in the present study are available upon reasonable request to the authors

---

**Supplementary Table 1:**
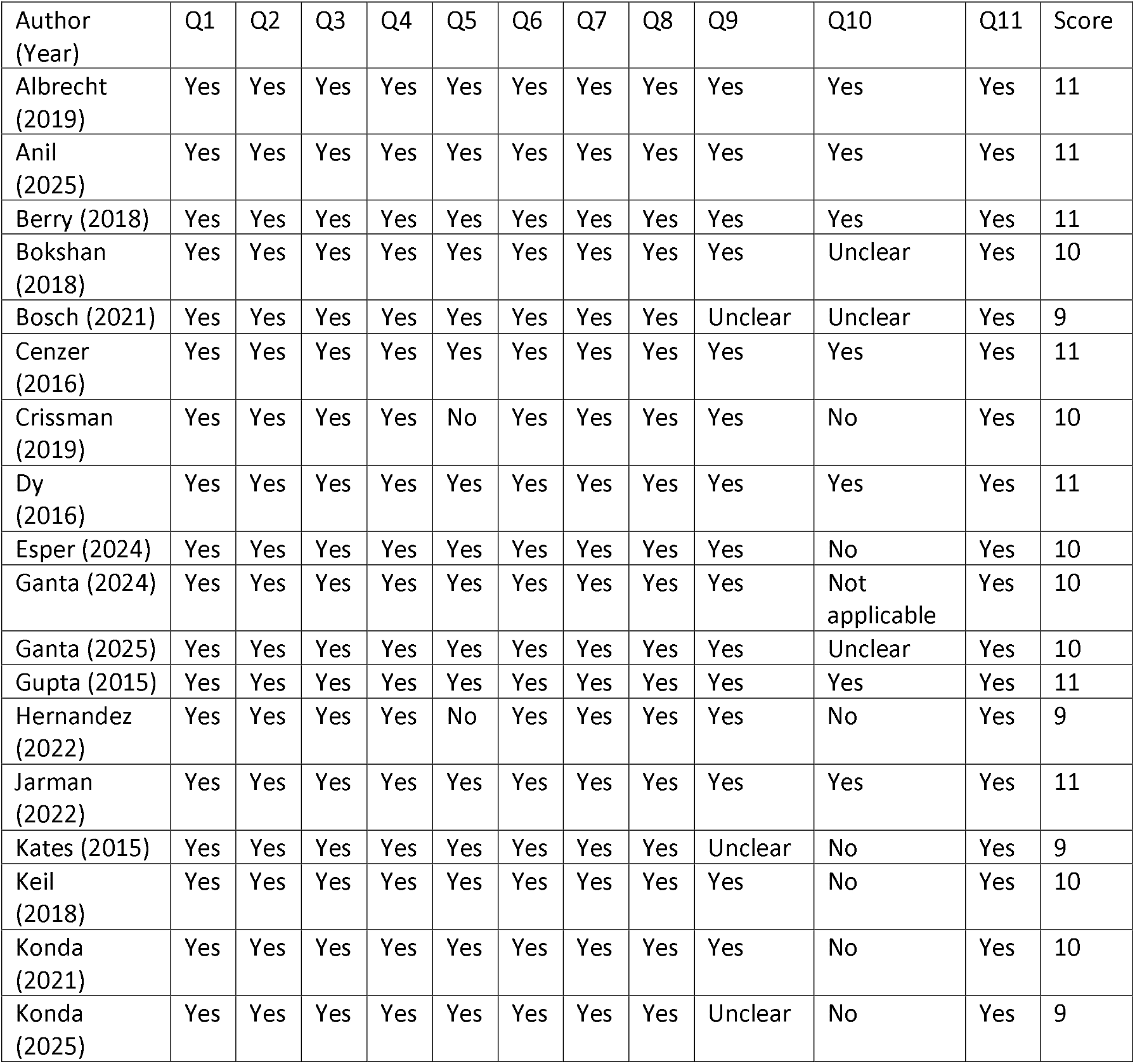
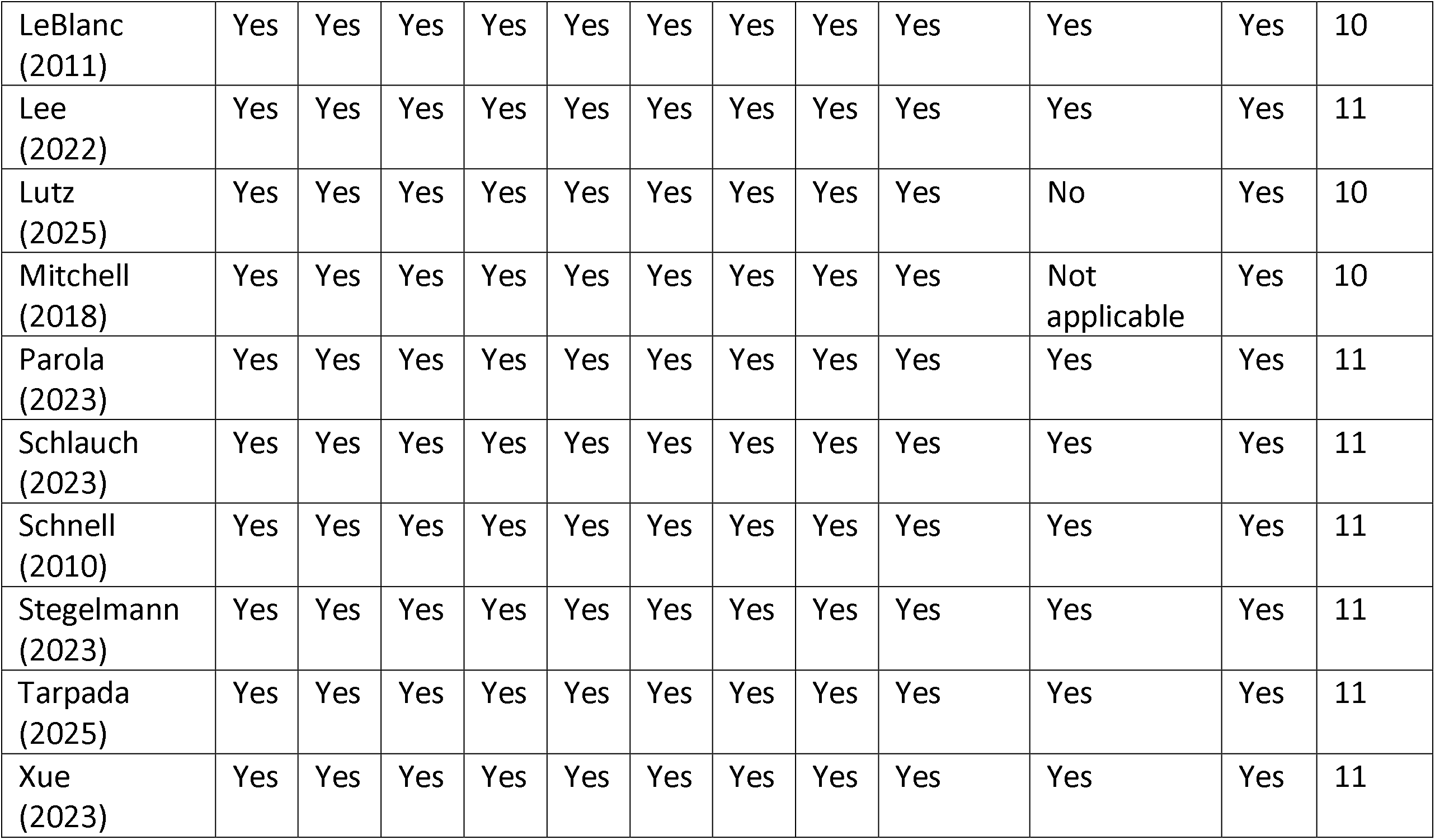
Joanna Briggs Institute for observational studies.

